# Manuscript title: Ultra-processed food consumption in UK adolescents: distribution, trends, and sociodemographic correlates using the National Diet and Nutrition Survey 2008/09 to 2018/19

**DOI:** 10.1101/2023.06.05.23290977

**Authors:** Yanaina Chavez-Ugalde, Frank De Vocht, Russell Jago, Jean Adams, Ken K. Ong, Nita Forouhi, Zoé Colombet, Luiza I.C. Ricardo, Esther Van Sluijs, Zoi Toumpakari

## Abstract

**Background:** The consumption of ultra-processed foods (UPF) has been proposed as a key driver of the global rise in non-communicable diseases. Evidence from several countries suggests that adolescents are the highest consumers. This study examined UPF consumption in a representative sample of UK adolescents.

**Methods:** We used data from 4-day food diaries from adolescents (11-18y) in the UK National Diet and Nutrition Survey (2008/09-2018/19) (n=3,270). UPF were identified using the NOVA classification. We estimated the percentage of Total Energy Intake (%TEI) and the absolute weight (grams). Linear regression models quantified differences in UPF consumption across survey years and its association with participant’s individual characteristics.

**Results:** Mean UPF consumption was 861 (SD 442) g/d and this accounted for 65.9% (SD 13.4%) of TEI. Between 2008 and 2019, mean UPF consumption decreased from 996 to 776 g/d [−211 (95%CI: −302;-120)] and from 67.7% to 62.8% of TEI [−4.8% (95%CI:-8.1;-1.5)]. Higher %TEI was consumed by adolescents with lower socioeconomic status; white ethnicity and living in England North. A higher weight of UPF consumption was associated with being male, white, age 18y, having parents with routine or manual occupation, living in England North, and living with obesity.

**Discussion and conclusion:** Average energy intake from UPF has decreased over a decade in UK adolescents. We observed a social and regional patterning of UPF consumption, with higher consumption among adolescents from lower socioeconomic backgrounds, from a white ethnicity and living in England North. Our findings suggest a relationship between individual characteristics and UPF consumption by UK adolescents.

## 1. Background

The consumption of ultra-processed foods (UPF) has been proposed as one of the key drivers of the global rise in chronic diet-related diseases (1). UPF are often manufactured from cheap industrial substances extracted or derived from foods, such as fats and oils, free sugars, and amino acids, which are then mixed with cosmetic additives, like colours, stabilisers, humectants, emulsifiers which are not used in domestic kitchens. UPF are often defined as nutritionally unhealthy due to their high content of sugar, salt and saturated fats (2). Some examples of UPF are soft drinks, breakfast cereals, reconstituted meat products, packaged breads, ready-to-eat foods. UPF are durable, convenient, ready-to-eat, hyper-palatable, have attractive packaging and are strongly marketed to children and adolescents (3). There is a growing body of evidence linking the consumption of UPF with poor dietary quality, and diet related diseases such as obesity in children, adolescents and adults (4), and chronic non-communicable diseases, such as cardiovascular disease and cancer (5) and all-cause mortality in adults (6).

Adolescence is a key transitional stage when major changes in practices that influence health occur (7). Additionally, adolescents’ search for novelty and openness to change makes them a vulnerable group for commercial marketing (3). Adolescents’ food patterns and practices are strongly driven by their food environments and eating contexts, their autonomy, peer influence and social norms (8). These can all be influenced by commercially targeted activities, such as marketing and advertising, product placement, and pricing strategies such as food promotions and discounts (9).

Evidence across different countries suggests that adolescents are the highest consumers on average of UPF compared to other age groups (10). In Canada it is estimated that UPF contribute approximately 55% of the total caloric intake in children and adolescents (2-18y) versus 45% in adults (11). In the USA, children (6-11y) (69.0%) and adolescents (12-19y) (67.7%) consumed a significantly higher percentage of energy from UPF than those aged 2-5 years (61.1%). (12).

Although consumption of UPF is becoming more prevalent worldwide (1, 13), there are notable cross-country and socioeconomic status (SES) differences (14). In nationally representative samples form high-income countries UPF contribute more than 50% of energy intake (15), and up to 30% in middle-income countries (16). Even though consumption of UPF is much higher in high-income compared to upper-middle income countries, within these latter countries adolescents are still the highest UPF consumption age group. Evidence from high- and upper-middle income countries signals a distinction in social patterning of UPF consumption according to the nutritional transition stage of each country (1).

Globally, the availability and sales of UPF have increased over time (2006–2019) (1), and evidence from the USA (2010–2018) and Korea (1999–2018) suggests that consumption of UPF among adolescents has also increased over time (12, 17). However, evidence from a representative sample of UK population (>1.5 years) did not find evidence for an increase in the energy share of UPF between 2008 and 2019 (18), however changes in weight consumption from UPF were not explored. Exploring UPF consumption over time could help us theorise behavioural patterns within a changing food environment. Given that adolescents are the highest consumers of UPF and evidence from other countries suggest that consumption in this age group has increased over time, investigating if and how UPF consumption in UK adolescents has changed over time remains an important question.

Using UPF as a descriptor of dietary patterns could potentially assist in the understanding of adolescents’ diet composition and inform the development of dietary guidelines and nutrition policy actions to improve diet quality and prevent diet-related chronic-diseases (14, 19).

Previous studies on UPF consumption have mostly expressed UPF as percentage of energy intake. However, the French NutriNet-Santé study (20) proposed expressing UPF by weight (grams) given that this can capture non-nutritional factors relating to processing of food and foods and drinks that contribute little to energy intake (e.g., additives, non-sugar sweeteners, neo-formed contaminants and endocrine-disrupting chemicals from packaging materials (21).

We used population-based individual level data from the UK to address the gaps in existing knowledge mentioned above. Specifically, the aims of this study were to: calculate UPF consumption and its contribution to relative energy intake (% kcal/day) and absolute food weight intake (g/day); describe UPF consumption across the 11 NDNS survey waves (2008–2019); and investigate if there are sociodemographic characteristics associated with UPF consumption in a representative sample of UK adolescents.

## 2. Methods

We conducted an analysis using repeat cross-sectional data from 11- to 18-year-old participants (adolescents) in the UK National Diet and Nutrition Survey Rolling Programme (NDNS) waves 1-11 (2008/09-2018/19). Data were downloaded from the UK Data Service (22). This study is reported according to the Strengthening the Reporting of Observational studies in Epidemiology – Nutritional Epidemiology (STROBE-nut). Parental consent was obtained for participants aged 11 to 15 and direct consent was obtained for participants aged 16 to 18 years. Additional ethical approval for this secondary analysis of anonymised data was not required.

### 2.1 Study design and population

The NDNS is an annual rolling programme (RP) of cross-sectional surveys conducted in the UK (England, Wales, Scotland, and Northern Ireland) assessing the food consumption, nutrient intake and nutritional status of the general UK population aged 1.5 years and above living in private households. NDNS survey year starts in April and data collection runs from April until March the following year. Population, sampling and recruitment are described in detail elsewhere (23). Briefly, NDNS’s continuous rolling programme seeks to collect data from a representative sample of the UK population with 1,000 participants every year (500 adults, 500 children) with provision of an additional sample to achieve country-level representativeness.

Sampling follows a multistage probability design to generate a new random sample of private households in the UK every year. Each year a random sample from small geographical areas [(i.e., primary sampling units (PSUs)] is selected. Within these PSUs, private addresses are randomly selected from the Postcode Address File and the selected households receive a visit by the study team. Within the participating household one child and one adult are randomly selected to take part. Participants within households are then asked to complete a food diary across 4 days to record all foods and beverages consumed inside and outside of the house. Those who record at least 3 days are then invited for further physical measurements.

#### Inclusion criteria

Individuals were included in the analysis if they took part in NDNS waves 1 to 11, were aged between ≥11 and ≤18 years at data collection and completed at least three out of four food diary days.

The current study combines NDNS data from waves 1 to 11 (2008/09 to 2018/19). Overall, at an individual level, 53% of those selected to take part, including adults and children, completed at least three food diary days. From the sample of included individuals, 3,270 were adolescents (11- to 18-year-olds).

### 2.2 Dietary data collection & processing

Dietary assessment in NDNS waves 1-11 was designed to provide full description, detail and quantification of all food and drink consumed during the dietary assessment period, with the ability to capture habitual consumption when conducted over a number of days. Seasonal variations in food and drink consumption are addressed by the continuous fieldwork design.

Trained interviewers collected sociodemographic information through interviews and administered the food diaries to adolescent participants. Adolescents were instructed (in the case of 11- and 12-year-olds, their parent or carer) by the trained interviewer to record location, time and quantity of all the food and drink they consumed inside and outside of the home over four consecutive days. Recording of the four-day food diary would start on selected days to ensure even representation of all days of the week across the whole sample. After the food diaries were completed, the interviewer returned to collect the diary, reviewed the data with the participant and added missing details to improve completeness. Participants received monetary gift vouchers following completion of at least three of the four dietary recordings. Household measures and nutritional information from labels were used to estimate portion sizes consumed. Food diaries were entered in the Dietary assessment system DINO (Diet In Nutrients Out) that uses food composition data from the NDNS Nutrient Databank (24, 25). Additional details of the coding of food intake data and calculation of energy intakes and food composition in NDNS data are described in detail elsewhere (24–27).

Across the 11 years of data collection, NDNS participants reported on 60 main food groups, 154 subsidiary food groups and 4,944 food items. Classification of foods and beverages according to their level of processing was conducted using the NOVA food classification system which considers the nature, extent and purpose of industrial food manufacturing processing (28). The NOVA classification includes four categories: 1) unprocessed or minimally processed foods, 2) processed culinary ingredients, 3) processed foods and 4) UPF. Figure 1 shows these four classifications and examples of each food group. More details on the NOVA classification can be found elsewhere (2, 28).

**Figure 1.**
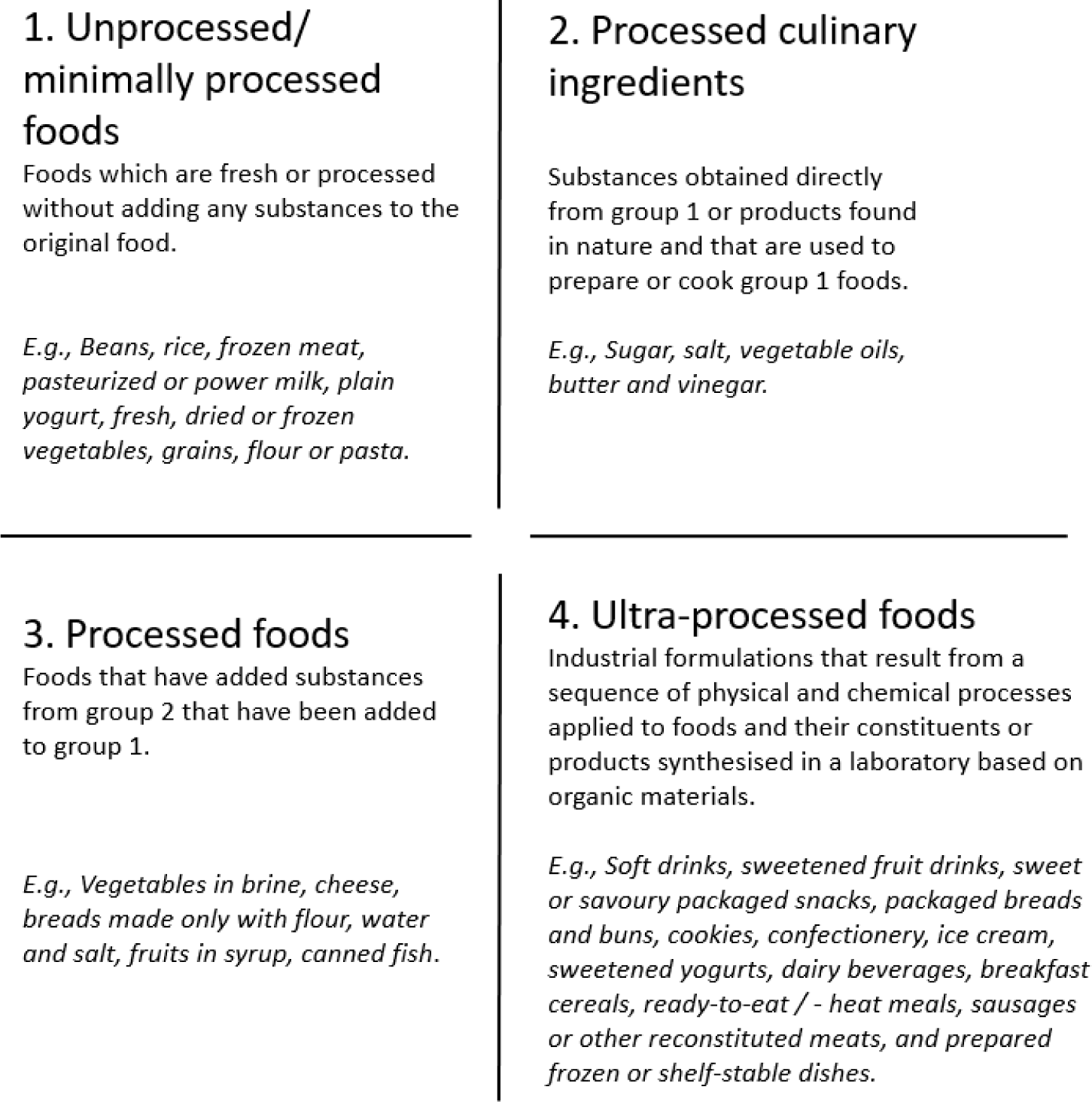
The 4 groups in the NOVA food classification system [based on information from (29)]

All foods and drinks in the NDNS Nutrient Databank were coded and grouped independently double-coded by two researchers (YCU and ZC) based on main food group first (e.g., ‘Pasta rice and other cereals’), then coded according to subsidiary food groups (e.g., pasta, manufactured products and ready meals) and then by individual food item (e.g., pasta, spaghetti, canned in tomato sauce) based on their recipe information and the ingredient list if the classification of a whole main or subgroup was not possible (e.g., composite dishes). This allowed us to classify each underlying or main constituent ingredient into the corresponding NOVA group as previously suggested on how to disaggregate composite food codes in NDNS data (26, 27). Food supplements (i.e., vitamins and minerals) were not coded. Previous studies that have used either the NOVA classification system and/or NDNS data served as a useful coding framework to classify dietary data in this study (4, 30, 31).

This study was specifically interested in the consumption of UPF. Thus, we classified group 1, 2 and 3 as non-UPF, and group 4 as UPF and focused on the UPF category (NOVA 4) to describe UPF consumption among UK adolescents.

#### NOVA classification level of agreement

The level of agreement between both coders was 97% after independently classifying all food and drink items using the full NOVA classification system (NOVA 1, 2, 3 and 4). Most of this disagreement (152 of 4,784 food and drink items, 3.1%) was between NOVA categories 1, 2 and 3. Only 10 of 4,784 (0.2%) food items caused a disagreement in UPF classification (NOVA 4). These 10 UPF were mainly composite dishes (i.e., beef curry takeaway, cottage pie with instant mash potato, black pudding in batter takeaway, fish pie with crust flaky pastry, vegetable pie with crust, *vol au vents* made with mushroom sauce and pastry), sauces (i.e., chilli pickle sweet, golden syrup), and ice-cream (i.e., ice-cream with double cream and purchased sorbet). These 10 food items categorised as NOVA 4 (UPF) by the first researcher (YCU) based on the individual food name and searches for lists of ingredients in branded products, whilst the second researcher (ZC) classified them based on the food name without searching for list of ingredients in branded items. Once the lists of ingredients were reviewed and discussed both researchers reached a 100% agreement.

### 2.3 Variables of interest

The outcomes of interest were the relative energy intake from UPF (NOVA 4) (% kcal/d) and the absolute weight of UPF consumed (g/d). These were calculated based on the relative energy and total absolute weight of foods and drinks classified as UPF in the NDNS food individual level dietary dataset reported by each participant. Individual-level sociodemographic characteristics included sex at birth, age, parent’s occupation social class, ethnic group and UK region. Other individual characteristics included weight categories derived from standardised body mass index (zBMI), survey year, and moderate-to-vigorous-physical activity (MVPA).

BMI was calculated from nurse measured height and weight. BMI values were standardised for sex and age and categorised [normal weight (<=1 standard deviation (sd)), overweight (>1 sd) and obese (>2 sd)] based on the 1990 British Growth Reference (UK90).

MVPA data was self-reported using the Recent Physical Activity Questionnaire (RPAQ) (32), which assessed type, amount of physical activity and screen time, and is validated for use in older adolescents and adults. Data was therefore collected only in 16–18-year-olds, and within these, only 56% provided MVPA data (18% of the entire sample, n= 575 out of n=3,270). Daily time spent in MVPA was summed in each individual (min/day). As MVPA was not normally distributed we created quartiles (<21 min/day; 21-52 min/day; 52-124 min/day; >124 min/day).

Other sociodemographic variables were categorised as follows: sex (male and female), age in completed whole years (8 categories from 11 to 18 years), parent’s occupation social class (higher managerial, administrative, and professional occupations; intermediate occupations; routine and manual occupations; we used the three level National Statistics Socioeconomic classification (NS-SEC)(33)), ethnic group (white, non-white), and region [England North, England Central/Midlands, England South (including London), Scotland, Wales, and Northern Ireland].

### 2.4 Statistical analysis

Study weights were provided by NDNS, and datasets were re-scaled and adjusted for the adolescent subsample based on NDNS study weights guide documentation (34). These weights were used using the *svy* prefix in Stata for all analysis to account for non-response and sampling error and to provide estimates representative of the UK adolescent population.

For the description of the sample, we reported weighted percentages (with 95%CIs) of the distribution of individual sociodemographic characteristics alongside the distribution and means of relative UPF energy consumption (%kcal/day) and absolute weight of UPF consumption (g/d) in the overall sample and for each sociodemographic characteristic. We display the mean of all available days of food diary for each individual.

We pooled all survey years and used multivariable linear regression models to test if dietary contribution of UPF (%kcal/d and g/d) differed across each of the variables of interest by doing individual stepwise models (i.e., sociodemographic categories and individual characteristics separately) adjusting for age, sex and survey year as covariates. Each category within a variable was compared to the reference group (i.e., sex (male), age (11 years) parent’s occupation social class (higher managerial, administrative, and professional occupations), ethnic group (white), and region (England North). We also used linear regression analysis to evaluate if mean UPF consumption (%kcal/d and g/d) changed across NDNS survey years by comparing each survey year (2 to 11) vs year 1 (2008/09) and adjusting for age and sex. Given the clustered regional sampling design of NDNS, we used survey analysis procedures to incorporate sample weights in these models.

#### Missing data

Variables with complete data (n=3,270) were sex, age, survey year, and region. Variables with missing data were ethnicity (0.1%), parents’ occupation (4.5%), BMI z-score (4.3%), and MVPA (82.4%). We used a complete case analysis for all analyses (n=2,991 for all variables, except for MVPA n=534, for which a separate analysis was done using this subsample).

##### Additional analysis

We ran an analysis adjusting for total energy intake in addition to the primary analysis to explore whether individual characteristics were associated with relative energy (%kcal/d) and absolute weight (g/d) independently from total energy intake.

All data analysis was conducted in Stata Statistical Software: Release 17.0 (StataCorp LP., College Station, TX, USA).

## 3. Results

Table 1 presents descriptive data of the 2,991participants with complete data for the variables of interest except MVPA (n=534). Of all adolescents, 51.3% were females, 42.9% had parents with a higher managerial, administrative and professional occupation, 66% had normal weight, 83.3% were from a white ethnic group, 43.7% lived in England South (including London), and of participants with physical activity data (16 to 18 year olds) 26.7% reported more than 124 min/day of MVPA.

**Table 1.** Descriptive characteristics of a weighted sample in the NDNS adolescents’ sample waves 1-11 (2008/09–2018/19) (n=2,991)

| Individual characteristics |  | %N (weighted)<sup>a</sup> | 95%CI |
| --- | --- | --- | --- |
| Sex | Male | 51.3 | (49.0, 53.6) |
|  | Female | 48.7 | (46.4, 51.0) |
| Age | 11 | 12.3 | (10.9, 13.9) |
|  | 12 | 12.8 | (11.3, 14.5) |
|  | 13 | 12.8 | (11.3, 14.5) |
|  | 14 | 12.6 | (11.1, 14.2) |
|  | 15 | 11.4 | (10.1, 12.8) |
|  | 16 | 14.6 | (13.0, 16.4) |
|  | 17 | 13.7 | (12.0, 15.5) |
|  | 18 | 9.9 | (8.6, 11.3) |
| Parent's occupation social class<sup>b</sup> | Higher managerial, administrative, and professional | 42.9 | (40.6, 45.3) |
|  | Intermediate | 22.8 | (20.9, 24.8) |
|  | Routine and manual | 34.3 | (32.1, 36.6) |
| Survey year<br>(survey wave for data collection) | (1) 2008/09 | 8.0 | (6.5, 9.9) |
|  | (2) 2009/10 | 10.1 | (8.2, 12.4) |
|  | (3) 2010/11 | 9.7 | (7.7, 12.1) |
|  | (4) 2011/12 | 8.4 | (6.8, 10.4) |
|  | (5) 2012/13 | 8.6 | (6.8, 10.9) |
|  | (6) 2013/14 | 9.2 | (7.4, 11.4) |
|  | (7) 2014/15 | 9.4 | (7.5, 11.7) |
|  | (8) 2015/16 | 8.6 | (6.8, 10.9) |
|  | (9) 2016/17 | 9.7 | (7.7, 12.2) |
|  | (10) 2017/18 | 9.0 | (7.1, 11.3) |
|  | (11) 2018/19 | 9.2 | (7.3, 11.5) |
| **Weight category<sup>c</sup>** | Normal weight | 66.0 | (63.6, 68.3) |
|  | Overweight | 13.7 | (12.1, 15.4) |
|  | Obese | 20.3 | (18.4, 22.4) |
| **Ethnic group** | White | 83.3 | (81.2, 85.1) |
|  | Non-white | 16.8 | (14.9, 18.8) |
| **Region** | England North | 22.9 | (21.2, 24.7) |
|  | England Central/Midlands | 17.7 | (16.1, 19.3) |
|  | England South (including London) | 43.7 | (41.8, 45.7) |
|  | Scotland | 7.7 | (6.4, 9.3) |
|  | Wales | 4.7 | (4.2, 5.3) |
|  | Northern Ireland | 3.2 | (2.9, 3.5) |
| **MVPA<sup>d</sup>** | <21 min/day | 26.9 | (21.9, 32.6) |
|  | 21-52 min/day | 22.4 | (18.0, 27.5) |
|  | 52 -124 min/day | 24.0 | (19.3, 29.4) |
|  | >124 min/day | 26.7 | (21.1, 33.2) |

<sup>a</sup> Percentages and means are weighed based on non-selection and non-response survey weights provided by NDNS year 2008-2019.
<sup>b</sup> Parents occupation is based on the three-class NS-SEC. A small number of households were excluded from this classification where the household has never worked or was classified under other.
<sup>c</sup> BMI z-score was created by standardising BMI for sex and age based on the 1990 British Growth Reference (UK90).
<sup>d</sup> MVPA: There was self-reported data available only for 16–18-year-olds, n=534.
**Abbreviations:** BMI z-score: standardised body mass index; MVPA: Moderate-to-vigorous physical activity; NDNS: National Diet and Nutrition Survey; NS-SEC: National Statistics Socioeconomic Class; UPF: ultra-processed food, NOVA4 classification group.

Table 2 shows the percentage of energy from UPF and grams per day of UPF consumed by adolescents by individual characteristics. Overall, adolescents reported a mean total energy intake per day of 1,741 kcal/day (SD 500.3) and 65.9% (SD 13.4%) of these calories come from UPF. In terms of food weight, adolescents consumed a mean of 2,004 (SD 727.3) g/day, of which 861 g/day (SD 442) was UPF (43% of total food weight consumed).

**Table 2.** Description of UPF consumption (%kcal and grams per day) of a weighted sample in the NDNS adolescents’ sample waves 1-11 (2008/09–2018/19)

|  |  | %<br>energy<br>from<br>UPF per<br>day<sup>a</sup> | 95%CI | Grams of<br>consumption<br>of UPF per<br>day<sup>a</sup> | 95%CI |
| --- | --- | --- | --- | --- | --- |
| **Sex** | Male | 66.0 | (65.0, 66.9) | 941.5 | (911.9, 971.2) |
|  | Female | 65.8 | (64.8, 66.7) | 776.3 | (748.6, 804.0) |
| **Age** | 11 | 65.6 | (63.9, 67.3) | 797.0 | (741.8, 852.3) |
|  | 12 | 66.1 | (64.3, 67.8) | 824.9 | (770.1, 879.7) |
|  | 13 | 68.0 | (66.1, 69.8) | 842.9 | (794.5, 891.4) |
|  | 14 | 67.2 | (65.5, 68.8) | 860.1 | (803.3, 916.9) |
|  | 15 | 66.0 | (64.4, 67.7) | 862.9 | (807.0, 918.8) |
|  | 16 | 66.0 | (64.4, 67.5) | 878.3 | (821.3, 935.2) |
|  | 17 | 64.4 | (62.1, 66.6) | 908.8 | (852.2, 965.4) |
|  | 18 | 63.4 | (61.3, 65.5) | 918.7 | (849.0, 988.3) |
| **Parents' occupation social class<sup>b</sup>** | Higher managerial, administrative, and professional | 63.8 | (62.8, 64.8) | 826.3 | (796.3, 856.2) |
|  | Intermediate | 65.9 | (64.6, 67.3) | 838.4 | (796.9, 879.9) |
|  | Routine and manual | 68.4 | (67.3, 69.6) | 919.6 | (883.4, 955.7) |
| **Survey year**<br>(survey wave for data collection) | (1) 2008/09 | 67.7 | (65.6, 69.8) | 995.5 | (928.1, 1,063.0) |
|  | (2) 2009-2010 | 68.1 | (66.2, 69.9) | 948.9 | (889.4, 1008.5) |
|  | (3) 2010-2011 | 67.9 | (66.4, 69.4) | 915.8 | (845.4, 986.2) |
|  | (4) 2011-2012 | 67.5 | (65.5, 69.5) | 880.8 | (814.9, 946.7) |
|  | (5) 2012-2013 | 67.0 | (65.0, 69.1) | 907.0 | (846.5, 967.6) |
|  | (6) 2013-2014 | 67.0 | (64.8, 69.2) | 876.7 | (816.1, 937.3) |
|  | (7) 2014-2015 | 67.3 | (65.4, 69.1) | 913.2 | (826.8, 999.5) |
|  | (8) 2015-2016 | 62.0 | (59.8, 64.2) | 746.0 | (680.8, 811.3) |
|  | (9) 2016-2017 | 62.8 | (60.2, 65.4) | 700.7 | (640.7, 760.6) |
|  | (10) 2017-2018 | 62.8 | (60.1, 65.4) | 820.7 | (745.6, 895.9) |
|  | (11) 2018-2019 | 64.6 | (62.1, 67.0) | 775.9 | (710.7, 841.0) |
| **Weight category<sup>c</sup>** | Normal weight | 65.8 | (64.9, 66.6) | 841.5 | (816.3, 866.8) |
|  | Overweight | 66.3 | (64.7, 67.9) | 861.4 | (810.8, 912.1) |
|  | Obese | 65.9 | (64.5, 67.4) | 924.1 | (873.0, 975.2) |
| **Ethnic group** | White | 67.3 | (66.6, 67.9) | 905.8 | (883.2, 928.5) |
|  | Non-white | 59.0 | (57.3, 60.8) | 638.4 | (596.4, 680.3) |
| **Region** | England North | 67.4 | (66.1, 68.8) | 910.8 | (866.6, 954.9) |
|  | England Central/Midlands | 66.8 | (65.3, 68.2) | 923.7 | (866.2, 981.3) |
|  | England South (including London) | 64.1 | (62.9, 65.2) | 803.9 | (771.2, 836.6) |
|  | Scotland | 67.8 | (65.7, 69.9) | 891.6 | (830.0, 953.2) |
|  | Wales | 67.1 | (65.5, 68.8) | 882.9 | (822.3, 943.5) |
|  | Northern Ireland | 67.9 | (66.6, 69.1) | 833.9 | (792.5, 875.3) |
| MVPA<sup>d</sup> | <21 min/day | 65.1 | (61.4, 68.8) | 865.1 | (776.1, 954.1) |
|  | 21-52 min/day | 65.5 | (62.7, 68.3) | 921.4 | (820.8, 1,022.0) |
|  | 52 -124 min/day | 65.3 | (62.1, 68.5) | 845.8 | (725.5, 966.2) |
|  | >124 min/day | 66.5 | (63.7, 69.3) | 1,052.2 | (962.7, 1,141.7) |

<sup>a</sup> Results are weighed based on non-selection and non-response survey weights provided by NDNS year 2008-2019.
<sup>b</sup> Parents occupation is based on the three-class NS-SEC. A small number of households were excluded from this classification where the household has never worked or was classified under other.
<sup>c</sup> BMI z-score was created by standardising BMI for sex and age based on the 1990 British Growth Reference (UK90).
<sup>d</sup> MVPA: There was data available only for 16–18-year-olds, n=534.
**Abbreviations:** BMI z-score: standardised body mass index; CI: Confidence Interval; MVPA: Moderate-to-vigorous physical activity; NDNS: National Diet and Nutrition Survey; NS-SEC: National Statistics Socioeconomic Class; UPF: ultra-processed food, NOVA4 classification group.

### Associations of UPF consumption with time and sociodemographic variables

#### UPF consumption across NDNS survey years

Figure 2 and Figure 3 (and Supplementary Table S1) show mean UPF consumption and CI’s across NDNS survey years (2009/09 – 2018/19) (adjusted for age and sex). There was evidence for a difference in relative energy (%kcal/day) and absolute weight (g/day) consumption from UPF across survey years (*p*<0.001).

**Figure 2.**
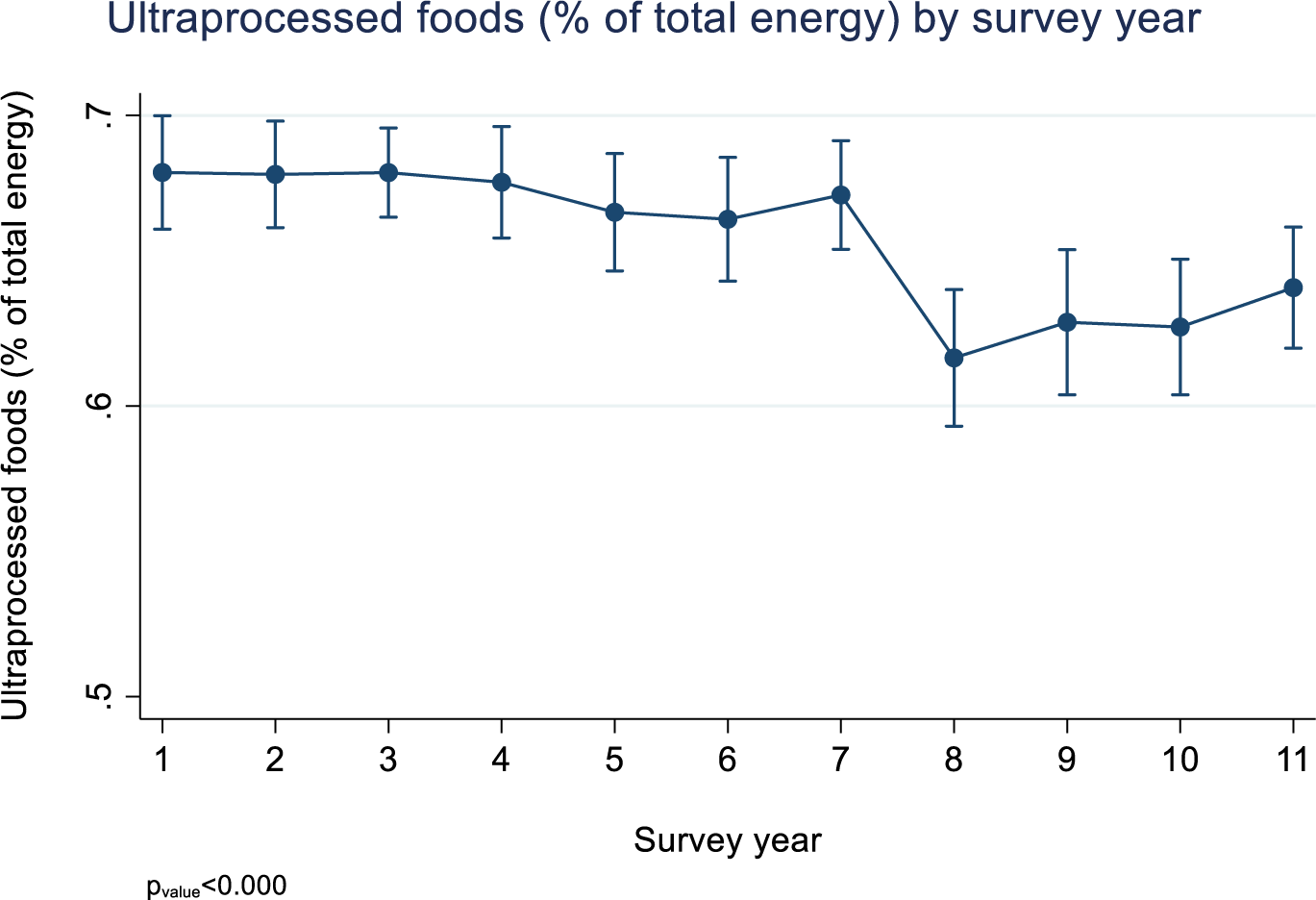
Mean relative energy consumption (%kcal/day) and CI’s from UPF across NDNS survey years (adjusted for age and sex)

**Figure 3.**
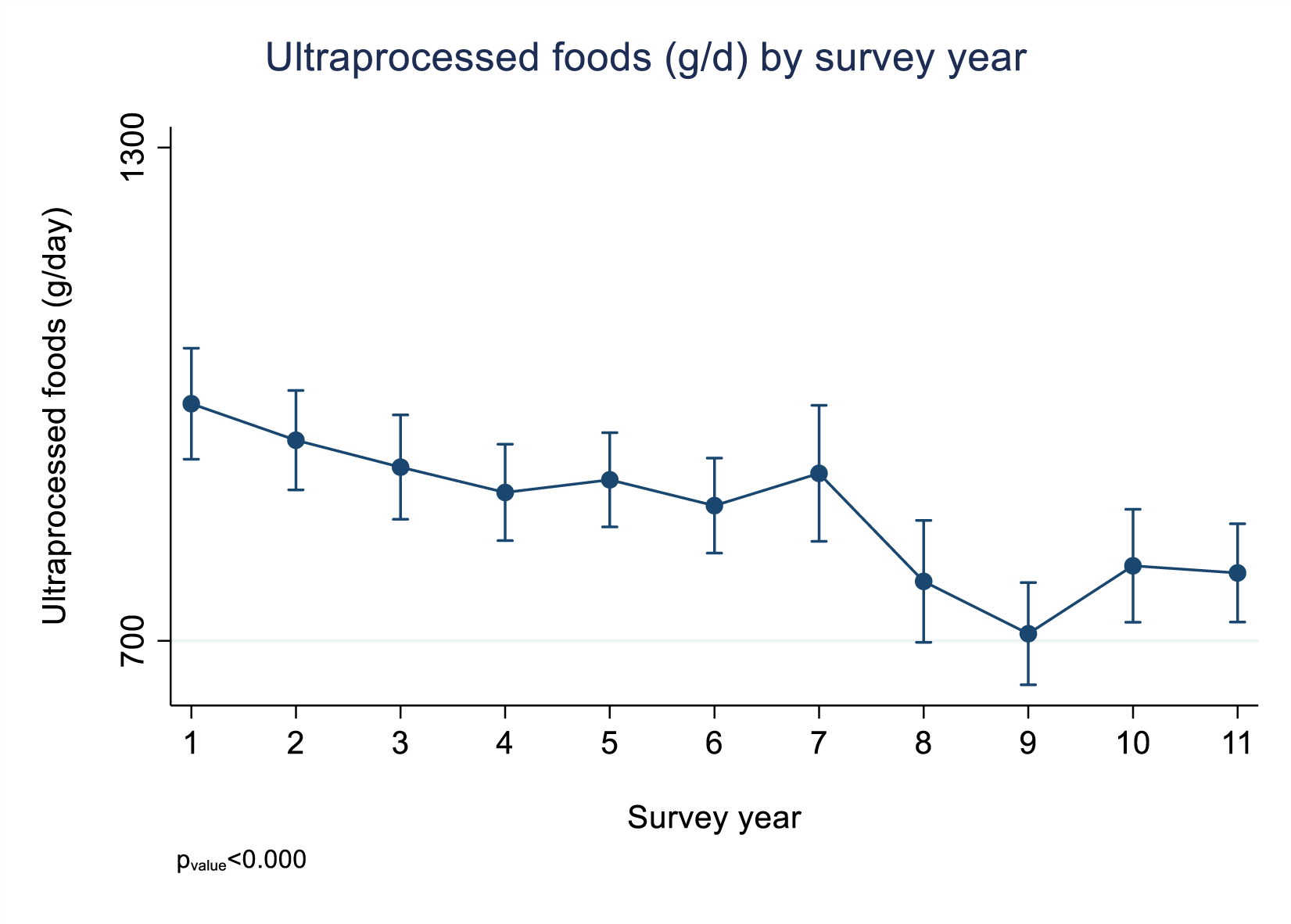
Mean grams of UPF consumption (plus 95% confidence interval) across NDNS survey years (adjusted for age and sex)

In Figure 2 we observed that comparing survey year 2 (2009/10) through 7 (2014/15) versus survey year 1 (i.e., reference year) there was no evidence of a difference in the absolute UPF as percentage of total energy intake (%TEI). However, %TEI from UPF was significantly lower from survey year 8 (2015/16) through 10 (2017/19) (vs year 1 – 2008/09) with the largest reduction in survey year 8 (−5.8 %kcal/day).

In Figure 3 we observed that the highest absolute weight from UPF consumed (g/d) was seen in year 1 (2008/09) (993.7 g/d). Similar to relative energy intake (%kcal/d), we observed a steady decline with some random variation in UPF weight across survey years.

#### Individual characteristics and %TEI from UPF

After adjusting for age, sex and survey year, results show (Figure 4 and Supplementary Table S1) that parents’ occupation, ethnic group and UK region were associated with percentage of energy consumption from UPF. A higher %TEI from UPF was consumed by adolescents whose parents had routine and manual occupations or intermediate occupations compared to adolescents with parents who had higher managerial occupations [intermediate: 2.0% (95%CI: 0.4; 3.5); routine and manual parental occupation: 4.6% (95%CI: 3.2; 6.1)]. Adolescents from a non-white ethnicity (vs white) reported lower %TEI from UPF [−8.0% (95%CI: −9.8; −6.1)] as well as those living in England South (including London) (vs England North) [−3.2% (95%CI: −4.9; −1.5)].

**Figure 4.**
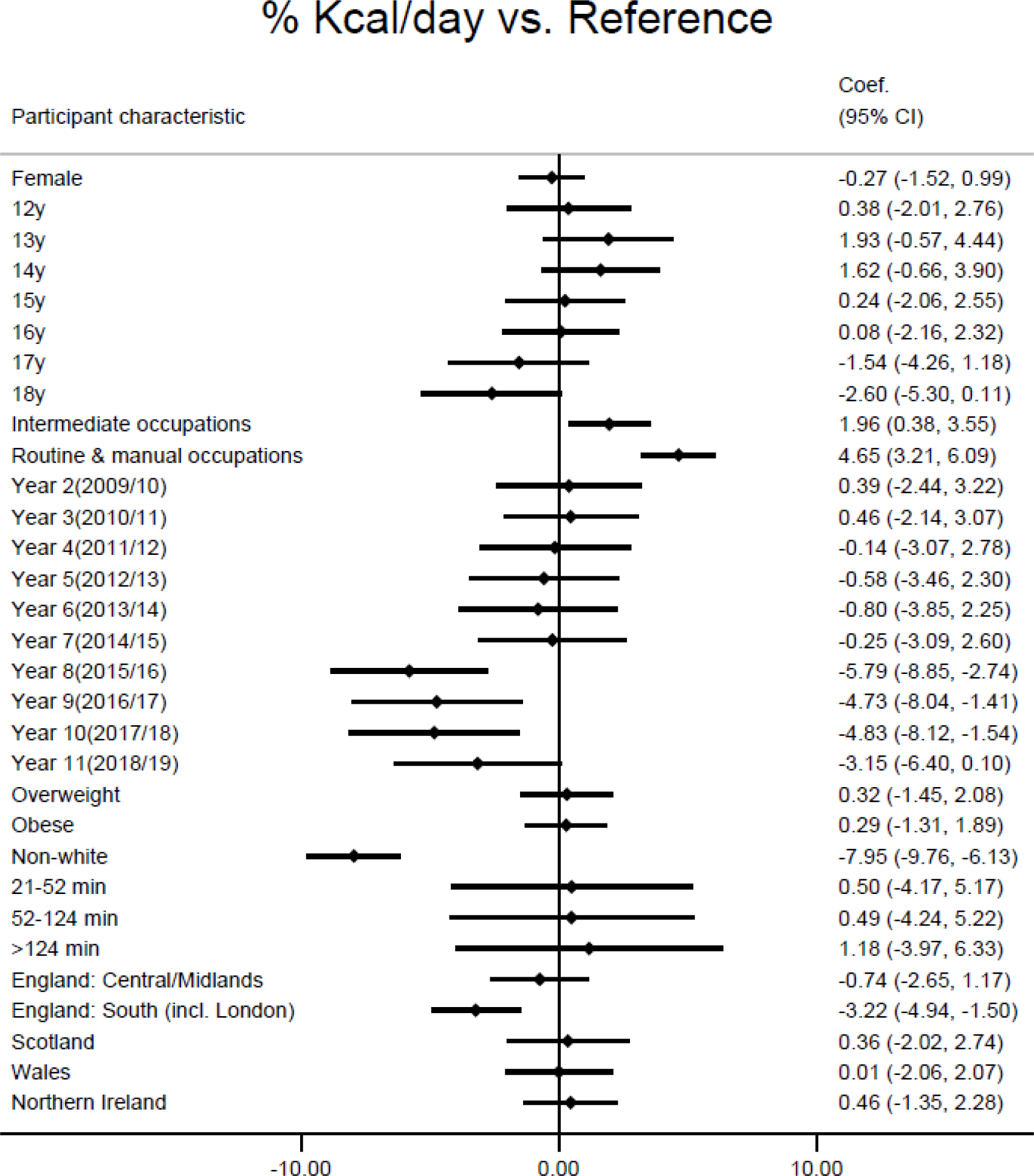
Linear regression of the adjusted associations between participants’ characteristics (vs reference category) and consumption of UPF defined as relative energy (%kcal/day) (adjusted for age, sex and survey year)

#### Individual characteristics and absolute weight from UPF

Figure 5 (and Supplementary Table S1) shows that higher weight of UPF consumption was associated with age between 17 and 18 years (vs 11-year-olds), with the highest consumption observed in 18-year-olds [115.1 g/day, (95%CI: 26.6; 203.5)]. Adolescents whose parents had a routine and manual occupation (vs higher managerial) [78.9 g/day, (95%CI: 34.6; 123.3)], and adolescents living with obesity (vs normal weight) [90.3 g/day (95%CI: 39.0; 141.5)] reported a higher UPF weight consumption. In contrast, lower weight of UPF consumption was associated with female sex (vs male) [−169.2 g/day, (95%CI: −206.8;-131.7)], non-white ethnicity (vs white) [−247.2, (95%CI: −292.8;-201.6)], and living in South England and Northern Ireland (vs. North England) [−99.2, (95%CI: −150.6;-47.8); −76.8, (95%CI: −135.0; −18.6)].

**Figure 5.**
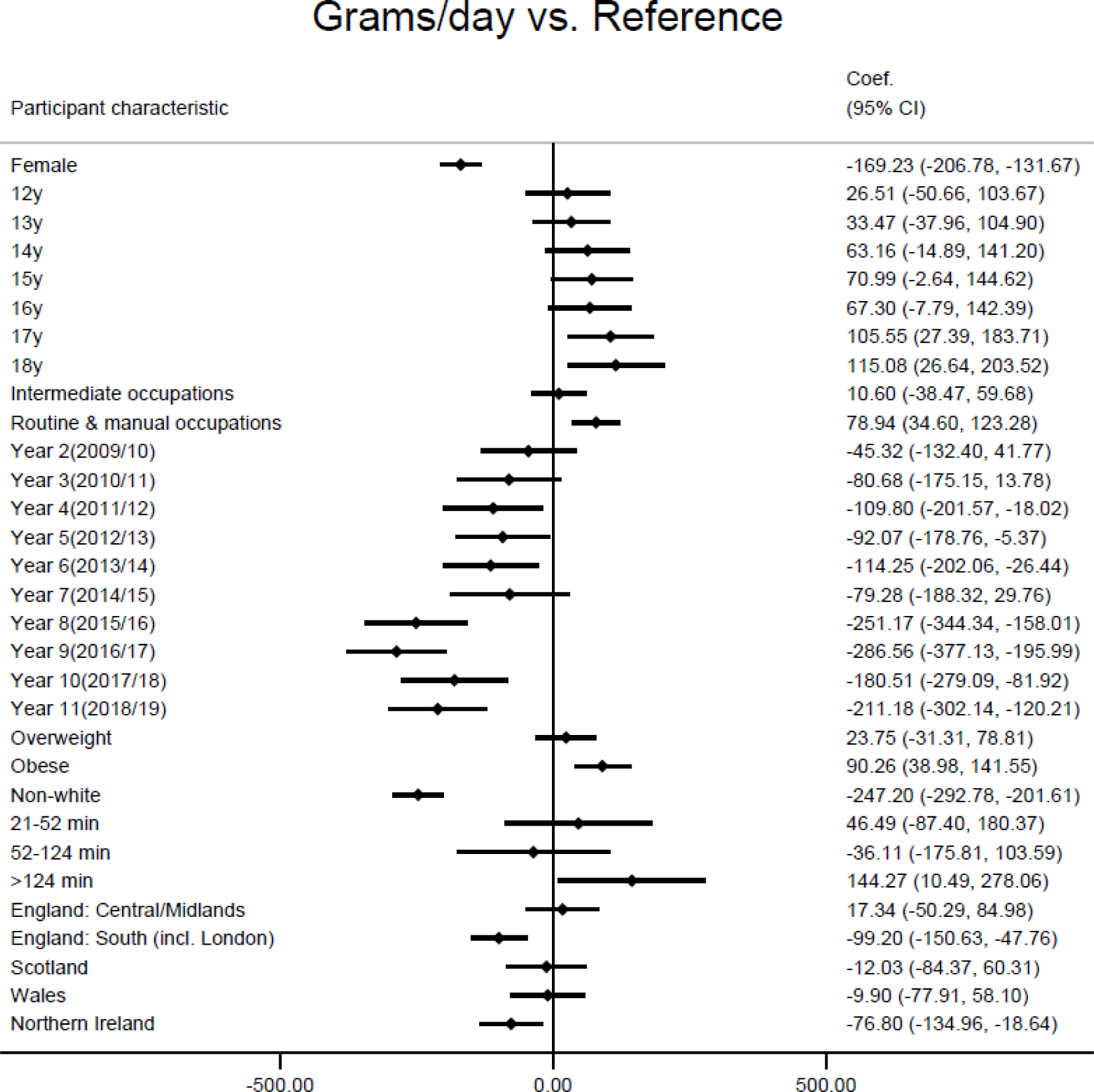
Linear regression of the adjusted associations between participants’ characteristics (vs reference category) and consumption of UPF defined as absolute weight (g/day)) (adjusted for age, sex and survey year)

#### Additional analysis

After additionally adjusting for total energy intake, all associations with %TEI from UPF persisted and in addition 18-year-olds (vs. 11-year-olds) had a lower %TEI (−3.0 %TEI, 95%CI: −5.7, −0.3). For UPF weight consumption, after adjusting for total energy intake associations were attenuated for age, survey years 4, 5, 6 and 10, and for MVPA >124 min/day (Supplementary Table S2).

## 4. Discussion

In this repeat cross-sectional study of a nationally representative sample of UK adolescents, we found that mean UPF among 11- to 18-year-olds was 861 g/day and accounted for 65.9% of their TEI. After adjusting for age and sex, mean consumption of UPF (both %TEI and weight) declined between 2008/09/ and 2018/19. Percentage of energy from UPF was lower by 3% and weight consumption from UPF was lower by 211.2 grams per day when comparing the first vs last NDNS survey waves (wave 1: 2008/09 vs wave 11:2018/19). After adjusting for age, sex and survey year, adolescents with lower socioeconomic status (having parents in an intermediate or routine and manual occupations), from a white ethnicity, and living in England North had higher %TEI from UPF. Additionally, being male, being between 17 and 18 years, having a lower socioeconomic status (having parents in a routine and manual occupation), living with obesity, from a white ethnicity, and living in England North were associated with a higher weight of UPF consumption. In additional analysis, adjusting for total energy intake, patterns of association with UPF weight became more consistent to those with %TEI from UPF.

These findings are consistent with previous analyses of the same NDNS data in 11 to 18-year-olds using survey years 2008/16 and 2008/17 (68% contribution of UPF to total energy intake (31, 35) and with analyses of data from adolescents living in other high-income countries (55% in Canada (11), and 68% in USA (12)). Although UPF are becoming more prevalent worldwide (1, 13, 14), there are cross-country differences that should be acknowledged. A recent multi-country study assessing adolescents’ UPF consumption in upper-middle and high-income countries found that consumption across these countries ranged between 19% to 36% in upper-middle-income countries (i.e., Argentina, Brazil, Colombia, and Mexico), and from 34% to 68% in high-income countries (i.e., Australia, Chile, USA and UK) with UK adolescents being the highest consumers of UPF (36). The high consumption of UPF within HIC such as the UK may be partly explained by a combination of social, cultural, economic, and marketing factors (14). Urbanisation in the UK, as in other HIC, can increase access to a greater diversity of and cheaper foods, including UPF, and increased exposure to commercial marketing with a wide offer of ready-to-eat products (37). These offer convenient solutions to longer working hours, changes in family structure and contribute to shifting from home food preparations to more ready-to-consume foods (38). An analysis of household availability of UPF in nineteen European countries showed that in the UK 50.4% of total purchased dietary energy comes from UPF, in contrast with 10.2% in Portugal, 13.4% in Italy and 46.2% in Germany (39).

Our findings suggest a decline of UPF consumption (%TEI and weight) between survey years 1 and 11 in the NDNS (2008/09 – 2018/19). Time trend analysis in NDNS (years 1 to 9) show a decline in total energy intake in this age group, especially between survey years 7 and 8 (2014/15). Additionally, the general downward trend in energy consumption from UPF is consistent with other study findings; a study using a controlled interrupted time series analysis between 2014 and 2019 found a 10% reduction in free sugars purchased per household per week from SSBs between 2014 and 2019 (40), however they did not find a reduction in volume (mL) purchased from SSBs. Another study using data from adults and children in NDNS found a reduction in energy share by 1.4% from sweetened beverages between 2008/09 to 2018/19, also without a reduction in volume (mL) consumed (18). This reduction could be partly explained by an increased public awareness and health concerns associated with sugar consumption, government-led campaigns and SSB reformulation to reduce sugar content. However, in this study we did not analyse UPF subgroup consumption and cannot attribute this drop in consumption exclusively to SSBs or free sugars. The reduction in UPF weight consumption in this study might be attributable to other UPF subgroups, or alternatively to a methodology change in NDNS dietary data collection.

Interestingly, in our sample UPF contributed proportionately more to TEI (65%) than to food weight (43%), which reflects the overall higher energy density of UPF. Energy density is associated with weight gain, type 2 diabetes, and obesity (41). Some associations were apparent with food weight but not with %TEI from UPF, for example with sex (higher in males vs. females) and age (higher in older adolescents). These differences largely reflect the higher TEI consumed by males and older adolescents. However, after accounting for TEI we still observed an independent significant effect. We need to understand mechanisms of harm, if any, to help guide finding a correct metric of exposure.

Similar to other findings, we observed that being from a white ethnicity is associated with a significantly higher consumption of UPF and less minimally processed foods (12, 35). However, the relationship between ethnicity and UPF is complex and multifactorial, and the observed higher UPF consumption among white ethnicity adolescents could be due to other factors associated with ethnicity, for example, cultural and other economic factors (42).

Our findings add to the body of knowledge that in HIC, lower socioeconomic groups consume higher levels of UPF (1). This may be partly due to the greater affordability (i.e., price per calorie) of less vs healthier foods across country incomes and regions (43) alongside targeted marketing to specific population subgroups (44). Additionally, reduced at-home facilities for food preparation and lack of cooking skills may lead to an increased consumption of more processed and convenience foods (e.g., pre-prepared meals) (45). Whilst in HIC most UPF are relatively cheaper than less processed foods, the opposite patterns are seen in LMIC. As an example, soft drinks are relatively inexpensive in HIC, whilst they are relatively expensive in LMIC (1, 14).

There is currently no universally agreed “safe” levels of dietary share from UPF. Therefore, future research to understand the mechanisms of harms to health may substantially improve nutritional quality of adolescent diets and contribute to the prevention of diet related NCDs.

### 4.1 Strengths and limitations

To the best of our knowledge, this is the first study to characterise and present data associating to both %TEI and weight of UPF consumption in a representative sample of UK adolescents.

This study has several additional strengths. Due to the consistent methods of data collection over time, the data across waves could be mixed resulting in a relatively large sample size. This study used individual-level dietary data, had 3 or 4 food diary days for each individual and had relatively high numbers of individuals within each group, which is likely to give a more accurate assessment of total dietary intake versus other methods (46, 47). For example, food records allow researchers to have high levels of detail on dietary intake, they were completed in real-time, which avoids reliance on recall, a common limitation of food frequency questionnaires and 24-hour recalls. Additionally, food records, when compared to direct observation and doubly labelled water, perform much better than other self-reported methods and capture about 80% of energy intake (46, 48). Weighting of the sample was applied to reduce non-response and sampling bias, therefore, the study results can be generalisable to the UK adolescent population. Additionally, this study included a weight measure of UPF to capture non-nutritional factors relating to processing of food (i.e., additives, non-sugar sweeteners, neo-formed contaminants), and foods and drinks that do not contribute to energy intake (e.g., artificially sweetened beverages). Our results show that there were more individual characteristics associated with weight of UPF consumption than UPF contribution to TEI. The inclusion of this measure could further enhance our knowledge about the risks involved by diets high in UPF beyond their contribution to TEI, but this should be systematically tested in studies of UPF and health outcomes.

To assess the variability and potential misclassification of UPF two researchers blindly classified all food and drink items in the food files within NDNS. We reached a 97% agreement in classification across all NOVA groups, and 99.8% agreement for classification of UPF. The variability of our classification of UPF showed that the energy contribution ranged from 65.9% under the current more conservative approach (YCU) to 70% when we applied the more liberal approach. Other studies that have assessed the variability and potential misclassification of UPF showed that <10% of individual foods and beverages reported in NHANES in the US (24-hr recall) were at risk of misclassification (49). This up to 4% variation range provides confidence in the current approach used to classify foods within NDNS according to the degree of processing using the NOVA classification system using food diaries.

Some limitations should be considered. We could not include a measure of household income due to the way this variable was collected in waves 9 to 11 limiting our knowledge of the impact of household income on UPF consumption among adolescents. However, other proxy for SES (i.e., parent’s social occupation social class) showed higher UPF consumption among lower SES groups.

Classification of food items according to the NOVA system was time-consuming because NDNS food diaries were not designed to capture UPF. Some of the main groups and subsidiary food groups were classified easily. However, composite food dishes had to be classified on an individual basis. Based on previous studies that have used either the NOVA classification system and/or NDNS data served as a practical coding framework (4, 5, 30, 31). Since the food records utilised were not designed to classify or evaluate foods according to their industrial processing, some items may have been misclassified. MVPA was self-reported was only available for a limited number of participants, which limited our ability to explore the associations of UPF and MVPA in a larger sample of adolescents.

## 5. Conclusion

This study showed that UK adolescents aged 11 to 18 years, living in England North, from the lowest SES group and with white ethnicity have the highest energy and weight intakes of UPF. Additionally, a higher weight consumption from UPF was observed in adolescents who were male, aged 17 to 18 years, and living with obesity. We found that the average energy intake and food weight from UPF has decreased in UK adolescents between year 1 and 11 of NDNS survey waves. However, it remains among the highest levels across high-income countries (e.g., Canada 55.0% and USA 67.7% of TEI). Estimating “safe” levels of dietary share from UPF and understanding the mechanisms of harms to health may substantially improve nutritional quality of adolescent diets and contribute to the prevention of diet-related NCDs.

## Data Availability

All data produced are available online at
https://beta.ukdataservice.ac.uk/datacatalogue/studies/study?id=6533
NOVA food classification are available upon reasonable request to the authors

https://beta.ukdataservice.ac.uk/datacatalogue/studies/study?id=6533

## Abbreviation list

CI: Confidence interval
HIC: High income country
LMIC: Low-middle-income country
MVPA: moderate-to-vigorous physical activity
NDNS: National Diet and Nutrition Survey
NS-SEC: National Statistics Socioeconomic Class
RPAQ: Recent physical activity questionnaire
TEI: Total energy intake
UK: United Kingdom
UPF: Ultra-processed food(s)
USA: United States of America
zBMI: Standardised Body mass index
ZC: Zoe Colombet

## Author contributions

YCU data collection, processing, design, data analysis, writing – first draft; FdV, ZT, RJ – conceptualisation, data processing; EVS, JA, KO, NF, ZT, FdV, RJ– editing, reviewing and supervision; ZC – coding and data processing; LICR – data visualisation. All authors have read and agreed to the final manuscript version.

## References

1. Baker P, Machado P, Santos T, Sievert K, Backholer K, Hadjikakou M, et al. Ultra-processed foods and the nutrition transition: Global, regional and national trends, food systems transformations and political economy drivers. Obesity reviews: an official journal of the International Association for the Study of Obesity. 2020;21(12):e13126.

2. Monteiro CA, Jaime PC, Cannon G, Levy RB, Louzada MLC, Rauber F, et al. Ultra-processed foods: What they are and how to identify them. Public Health Nutrition. 2019;22(5):936–41.

3. Norman J, Kelly B, Boyland E, McMahon A-T. The Impact of Marketing and Advertising on Food Behaviours: Evaluating the Evidence for a Causal Relationship. Current Nutrition Reports. 2016;5(3):139–49.

4. Chang K, Khandpur N, Neri D, Touvier M, Huybrechts I, Millett C, et al. Association Between Childhood Consumption of Ultraprocessed Food and Adiposity Trajectories in the Avon Longitudinal Study of Parents and Children Birth Cohort. JAMA pediatrics. 2021;175(9):e211573.

5. 5. Rauber F, da Costa Louzada ML, Steele EM, Millett C, Monteiro CA, Levy RB. Ultra-Processed Food Consumption and Chronic Non-Communicable Diseases-Related Dietary Nutrient Profile in the UK (2008⁻2014). Nutrients. 2018;10(5).

6. Kim H, Hu EA, Rebholz CM. Ultra-processed food intake and mortality in the USA: results from the Third National Health and Nutrition Examination Survey (NHANES III, 1988-1994). Public health nutrition. 2019;22(10):1777–85.

7. Olstad DL, Kirkpatrick SI. Planting seeds of change: reconceptualizing what people eat as eating practices and patterns. International Journal of Behavioral Nutrition and Physical Activity. 2021;18(1):32.

8. Patton GC, Sawyer SM, Santelli JS, Ross DA, Afifi R, Allen NB, et al. Our future: a Lancet commission on adolescent health and wellbeing. Lancet (London, England). 2016;387(10036):2423–78.

9. Chavez-Ugalde Y, Jago R, Toumpakari Z, Egan M, Cummins S, White M, et al. Conceptualising the commercial determinants of dietary behaviors associated with obesity: A systematic review using principles from critical interpretative synthesis. Obesity science & practice. 2021;7(4):473–86.

10. Leonie E, Machado P, Zinöcker M, Baker P, Lawrence M. Ultra-Processed Foods and Health Outcomes: A Narrative Review. Nutrients. 2020;12(7).

11. Moubarac J-C, Batal M, Louzada ML, Martinez Steele E, Monteiro CA. Consumption of ultra-processed foods predicts diet quality in Canada. Appetite. 2017;108:512–20.

12. Wang L, Martínez Steele E, Du M, Pomeranz JL, O’Connor LE, Herrick KA, et al. Trends in Consumption of Ultraprocessed Foods Among US Youths Aged 2-19 Years, 1999-2018. JAMA. 2021;326(6):519–30.

13. Monteiro CA, Moubarac JC, Cannon G, Ng SW, Popkin B. Ultra-processed products are becoming dominant in the global food system. Obesity Reviews. 2013;14(S2):21–8.

14. Moodie R, Bennett E, Kwong EJL, Santos TM, Pratiwi L, Williams J, et al. Ultra-Processed Profits: The Political Economy of Countering the Global Spread of Ultra-Processed Foods - A Synthesis Review on the Market and Political Practices of Transnational Food Corporations and Strategic Public Health Responses. Int J Health Policy Manag. 2021;10(12):968–82.

15. Martínez Steele E, Baraldi LG, Louzada ML, Moubarac JC, Mozaffarian D, Monteiro CA. Ultra-processed foods and added sugars in the US diet: evidence from a nationally representative cross-sectional study. BMJ Open. 2016;6(3):e009892.

16. Marrón-Ponce JA, Sánchez-Pimienta TG, Louzada M, Batis C. Energy contribution of NOVA food groups and sociodemographic determinants of ultra-processed food consumption in the Mexican population. Public Health Nutr. 2018;21(1):87–93.

17. Shim JS, Shim SY, Cha HJ, Kim J, Kim HC. Socioeconomic Characteristics and Trends in the Consumption of Ultra-Processed Foods in Korea from 2010 to 2018. Nutrients. 2021;13(4).

18. Madruga M, Martínez Steele E, Reynolds C, Levy RB, Rauber F. Trends in food consumption according to the degree of food processing among the UK population over 11 years. Br J Nutr. 2022:1–8.

19. Neufeld LMP, Andrade EBP, Ballonoff Suleiman AD, Barker MP, Beal TP, Blum LSP, et al. Food choice in transition: adolescent autonomy, agency, and the food environment. The Lancet. 2022;399(10320):185–97.

20. Julia C, Martinez L, Allès B, Touvier M, Hercberg S, Méjean C, et al. Contribution of ultra-processed foods in the diet of adults from the French NutriNet-Santé study. Public Health Nutr. 2018;21(1):27–37.

21. Lustig RH, Collier D, Kassotis C, Roepke TA, Kim MJ, Blanc E, et al. Obesity I: Overview and molecular and biochemical mechanisms. Biochemical Pharmacology. 2022;199:115012.

22. National Diet and Nutrition Survey Years 1-8, 2008/09-2015/16. [data collection] [Internet]. UK Data Service. 2018. Available from: https://beta.ukdataservice.ac.uk/datacatalogue/studies/study?id=6533&type=Data%20catalogue#!/details.

23. Public Health England. Appendix A. Dietary data collection and editing. In National Diet and Nutrition Survey. Results from Years 1–4 (Combined) of the Rolling Programme (2008/2009– 2011/2012). London, UK: Public Health England; 2014.

24. Fitt E, Cole D, Ziauddeen N, Pell D, Stickley E, Harvey A, et al. DINO (Diet in Nutrients Out)-An integrated dietary assessment system. Public Health Nutrition. 2015;18(2):234–41.

25. Public Health England. McCance and Widdowson’s the composition of foods integrated dataset 2015. In: Public Health England, editor. London, UK 2015.

26. Fitt E, Mak TN, Stephen AM, Prynne C, Roberts C, Swan G, et al. Disaggregating composite food codes in the UK National Diet and Nutrition Survey food composition databank. European journal of clinical nutrition. 2010;64(SUPP/3):S32–S6.

27. MRC Human Nutrition Research. Food Standards Agency Standard Recipes Database, 1992-2012 [data collection]. In: UK Data Service, editor. 2017.

28. Monteiro CA, Jaime PC, Cannon G, Moubarac JC, Levy RB, Louzada MLC. The UN Decade of Nutrition, the NOVA food classification and the trouble with ultra-processing. Public Health Nutrition. 2018;21(1):5–17.

29. Monteiro CA, Cannon, G., Lawrence, M., Costa Louzada, M.L. and Pereira Machado, P. Ultra-processed foods, diet quality, and health using the NOVA classification system. Rome, FAO. 2019.

30. Martines RM, Machado PP, Neri DA, Levy RB, Rauber F. Association between watching TV whilst eating and children’s consumption of ultraprocessed foods in United Kingdom. Maternal and child nutrition. 2019;15(4):n/a-n/a.

31. Rauber F, Martins CA, Azeredo CM, Leffa PS, Louzada MLC, Levy RB. Eating context and ultraprocessed food consumption among UK adolescents. The British journal of nutrition. 2021:1–11.

32. Golubic R, May AM, Benjaminsen Borch K, Overvad K, Charles M-A, Diaz MJT, et al. Validity of Electronically Administered Recent Physical Activity Questionnaire (RPAQ) in Ten European Countries. PLoS ONE. 2014;9(3):e92829.

33. Rose D., Pevalin D., K. OR. The National Statistics Socio-economic Classification: origins, development and use: Palgrave Macmillan UK; 2005.

34. UK Data Archive Study. Weights Guide: Combining data from Years 1-4, Years 5&6, Years 7&8 and Years 9-11, National Diet and Nutrition Survey. 2020.

35. Parnham JC, Chang K, Rauber F, Levy RB, Millett C, Laverty AA, et al. The Ultra-Processed Food Content of School Meals and Packed Lunches in the United Kingdom. Nutrients. 2022;14(14).

36. Neri D, Steele EM, Khandpur N, Cediel G, Zapata ME, Rauber F, et al. Ultraprocessed food consumption and dietary nutrient profiles associated with obesity: A multicountry study of children and adolescents. Obesity Reviews. 2022;23(S1).

37. Popkin BM, Adair LS, Ng SW. Global nutrition transition and the pandemic of obesity in developing countries. Nutrition Reviews. 2012;70(1):3–21.

38. Smith LP, Ng SW, Popkin BM. Trends in US home food preparation and consumption: analysis of national nutrition surveys and time use studies from 1965-1966 to 2007-2008. Nutr J. 2013;12:45.

39. Monteiro CA, Moubarac JC, Levy RB, Canella DS, Louzada M, Cannon G. Household availability of ultra-processed foods and obesity in nineteen European countries. Public Health Nutr. 2018;21(1):18–26.

40. Pell D, Mytton O, Penney TL, Briggs A, Cummins S, Penn-Jones C, et al. Changes in soft drinks purchased by British households associated with the UK soft drinks industry levy: controlled interrupted time series analysis. BMJ. 2021;372:n254.

41. Wang J, Luben R, Khaw K-T, Bingham S, Wareham NJ, Forouhi NG. Dietary Energy Density Predicts the Risk of Incident Type 2 Diabetes: The European Prospective Investigation of Cancer (EPIC)-Norfolk Study. Diabetes Care. 2008;31(11):2120–5.

42. Poti JM, Mendez MA, Ng SW, Popkin BM. Is the degree of food processing and convenience linked with the nutritional quality of foods purchased by US households? Am J Clin Nutr. 2015;101(6):1251–62.

43. Headey DD, Alderman HH. The Relative Caloric Prices of Healthy and Unhealthy Foods Differ Systematically across Income Levels and Continents. J Nutr. 2019;149(11):2020–33.

44. Kelly B, Vandevijvere S, Freeman B, Jenkin G. New Media but Same Old Tricks: Food Marketing to Children in the Digital Age. Curr Obes Rep. 2015;4(1):37–45.

45. Adams J, White M. Characterisation of UK diets according to degree of food processing and associations with socio-demographics and obesity: cross-sectional analysis of UK National Diet and Nutrition Survey (2008–12). International Journal of Behavioral Nutrition and Physical Activity. 2015;12(1):160.

46. Black AE, Prentice AM, Goldberg GR, Jebb SA, Bingham SA, Livingstone MB, et al. Measurements of total energy expenditure provide insights into the validity of dietary measurements of energy intake. Journal of the American Dietetic Association. 1993;93(5):572–9.

47. Bingham SA. Limitations of the Various Methods for Collecting Dietary Intake Data. Annals of Nutrition and Metabolism. 1991;35(3):117–27.

48. Park Y, Dodd KW, Kipnis V, Thompson FE, Potischman N, Schoeller DA, et al. Comparison of self-reported dietary intakes from the Automated Self-Administered 24-h recall, 4-d food records, and food-frequency questionnaires against recovery biomarkers. The American journal of clinical nutrition. 2018;107(1):80–93.

49. Steele EM, O’Connor LE, Juul F, Khandpur N, Galastri Baraldi L, Monteiro CA, et al. Identifying and Estimating Ultraprocessed Food Intake in the US NHANES According to the Nova Classification System of Food Processing. The Journal of Nutrition. 2023;153(1):225–41.

