## Supplementary material (Table S1 and S2) for "Manuscript title: Ultra-processed food consumption in UK adolescents: distribution, trends, and sociodemographic correlates using the National Diet and Nutrition Survey 2008/09 to 2018/19"

Supplementary Table S1 Associations between participants’ individual characteristics and consumption of UPFs defined as percentage of energy and grams per day in a weighted sample in NDNS adolescents (11 to 18 year-olds) sample waves 1-11 (2008/09–2018/19)

|  | | | % energy from UPF per day^a^ | | | | Grams of consumption of UPF per day^a^ | | |
| --- | --- | --- | --- | --- | --- | --- | --- | --- | --- |
| Characteristic | | | Adjusted coefficient* | | 95% CI | | Adjusted coefficient* | | 95% CI |
| Sex^I^ | Male | Reference | |  | | Reference | |  | |
|  | Female | -0.27 | | (-1.5, 1.0) | | **-169.2** | | **(-206.8, -131.7)** | |
| Age^II^ | 11 | Reference | |  | | Reference | |  | |
|  | 12 | 3.8 | | (-2.0, 2.8) | | 26.5 | | (-50.7, 103.7) | |
|  | 13 | 1.9 | | (-0.6, 4.4) | | 33.5 | | (-38.0, 104.9) | |
|  | 14 | 1.6 | | (-0.7, 3.9) | | 63.2 | | (-14.9, 141.2) | |
|  | 15 | 0.2 | | (-2.1, 2.5) | | 71.0 | | (-2.6, 144.6) | |
|  | 16 | 0.1 | | (-2.2, 2.3) | | 67.3 | | (-7.8, 142.4) | |
|  | 17 | -1.5 | | (-4.3, 1.2) | | **105.6** | | **(27.4, 183.7)** | |
|  | 18 | -2.6 | | (-5.3, 0.1) | | **115.1** | | **(26.6, 203.5)** | |
| Parent’s occupation social class^b^ | Higher managerial, administrative, and professional | Reference | |  | | Reference | |  | |
|  | Intermediate | **2.0** | | **(0.4, 3.5)** | | 10.6 | | (-38.5, 59.7) | |
|  | Routine and manual | **4.6** | | **(3.2, 6.1)** | | **78.9** | | **(34.6, 123.3)** | |
| Survey year ^III^  (survey wave for data collection) | (1) 2008/09 | Reference | |  | | Reference | |  | |
|  | (2) 2009/10 | 0.4 | | (-2.4, 3.2) | | -45.3 | | (-132.4, 41.8) | |
|  | (3) 2010/11 | 0.5 | | (-2.1, 3.1) | | -80.7 | | (-175.1, 13.8) | |
|  | (4) 2011/12 | -0.1 | | (-3.1, 2.8) | | **-109.8** | | **(-201.6, -18.0)** | |
|  | (5) 2012/13 | -0.6 | | (-3.5, 2.3) | | **-92.1** | | **(-178.8, -5.4)** | |
|  | (6) 2013/14 | -0.8 | | (-3.8, 2.3) | | **-114.2** | | **(-202.1, -26.4)** | |
|  | (7) 2014/15 | -0.2 | | (-3.1, 2.6) | | -79.3 | | (-188.3, 29.8) | |
|  | (8) 2015/16 | **-5.8** | | **(-8.8, -2.7)** | | **-251.2** | | **(-344.3, -158.0)** | |
|  | (9) 2016/17 | **-4.7** | | **(-8.0, -1.4)** | | **-286.6** | | **(-377.1, -196.0)** | |
|  | (10) 2017/18 | **-4.8** | | **(-8.1, -1.5)** | | **-180.5** | | **(-279.1, -81.9)** | |
|  | (11) 2018/19 | -3.2 | | (-6.4, 0.1) | | **-211.2** | | **(-302.1, -120.2)** | |
| Weight category^c^ | Normal weight | Reference | |  | | Reference | |  | |
|  | Overweight | 0.3 | | (-1.5, 2.1) | | 23.8 | | (-31.3, 78.8) | |
|  | Obese | 0.3 | | (-1.3, 1.9) | | **90.3** | | **(39.0, 141.5)** | |
| Ethnic group^d^ | White | Reference | |  | | Reference | |  | |
|  | Non-white | **-8.0** | | **(-9.8, -6.1)** | | **-247.2** | | **(-292.8, -201.6)** | |
| Region | England North | Reference | |  | | Reference | |  | |
|  | England Central/Midlands | -0.7 | | (-2.7, 1.2) | | 17.3 | | (-50.3, 85.0) | |
|  | England South (including London) | **-3.2** | | **(-4.9, -1.5)** | | **-99.2** | | **(-150.6, -47.8)** | |
|  | Scotland | -0.4 | | (-2.0, 2.7) | | -12.0 | | (-84.4, 60.3) | |
|  | Wales | 0.0 | | (-2.1, 2.1) | | -9.9 | | (-77.9, 58.1) | |
|  | Northern Ireland | 0.5 | | (-1.3, 2.3) | | **-76.8** | | **(-135.0, -18.6)** | |
| MVPA^e^ | <21 min/day | Reference | |  | | Reference | |  | |
|  | 21-52 min/day | 0.5 | | (-4.2, 5.2) | | 46.5 | | (-87.4, 180.4) | |
|  | 52 -124 min/day | 0.5 | | (-4.2, 5.2) | | -36.1 | | (-175.8, 103.6) | |
|  | >124 min/day | 1.2 | | (-4.0, 6.3) | | **144.3** | | **(10.5, 278.1)** | |
| ^*^ Sample size with complete data on variables of interest n=2,991  *Adjusted for Age, Sex and Survey Year.  ^I^Only adjusted for Age and Survey Year when Sex was the exposure  ^II^***Only adjusted for Sex and Survey Year when Age was the exposure  ^III^ Only adjusted for Sex and Age when Survey year was the exposure  ^a^ Percentages and means are weighed based on non-selection and non-response survey weights provided by NDNS year 2008-2019.  ^b^ Parents occupation is based on the three-class NS-SEC. A small number of households were excluded from this classification where the household has never worked or was classified under other.  ^c^ Weight category = BMI z-score was created by standardising BMI for sex and age based on the 1990 British Growth Reference (UK90).  ^d^ 0.1% missing data for ethnic group.  ^e^ MVPA: (n=534) There was 82.4% of missing data for MVPA. There was data available only for 16–18-year-olds and within these, there was 54.2% of missing data  Abbreviations: BMI z-score: standardised body mass index; MVPA: Moderate-to-vigorous physical activity; NDNS: National Diet and Nutrition Survey; NS-SEC: National Statistics Socioeconomic Class; UPF: ultra-processed food, NOVA4 classification group. | | | | | | | | | |

Supplementary Table S2 Additional analysis adjusting for total energy intake, age, sex and survey year. Associations between participants’ individual characteristics and consumption of UPFs defined as percentage of energy and grams per day in a weighted sample in NDNS adolescents (11 to 18 year-olds) sample waves 1-11 (2008/09–2018/19) (n=2,991)

|  | | | % energy from UPF per day^a^ | | | | Grams of consumption of UPF per day^a^ | | |
| --- | --- | --- | --- | --- | --- | --- | --- | --- | --- |
| Characteristic | | | **Adjusted coefficient*** | | **95% CI** | | **Adjusted coefficient*** | | **95% CI** |
| Sex** | Male | Reference | |  | | Reference | |  | |
|  | Female | 0.47 | | (-0.9, 1.8) | | **-37.3** | | **(-72.8, -1.8)** | |
| Age*** | 11 | Reference | |  | | Reference | |  | |
|  | 12 | 2.3 | | (-2.1, 2.6) | | -0.2 | | (-69.1, 68.8) | |
|  | 13 | 1.8 | | (-0.7, 4.3) | | 11.2 | | (-56.6, 78.9) | |
|  | 14 | 1.4 | | (-0.8, 3.7) | | 27.6 | | (-43.5, 98.7) | |
|  | 15 | -0.1 | | (-2.4, 2.2) | | 7.1 | | (-62.2, 76.4) | |
|  | 16 | -0.3 | | (-2.5, 2.0) | | 5.7 | | (-62.5, 73.9) | |
|  | 17 | -1.9 | | (-4.6, 0.9) | | 47.7 | | (-25.2, 120.7) | |
|  | 18 | **-3.0** | | **(-5.7, -0.3)** | | 44.4 | | (-37.2, 126.0) | |
| Parent’s occupation social class^b^ | Higher managerial, administrative, and professional | Reference | |  | | Reference | |  | |
|  | Intermediate | **2.1** | | **(0.5, 3.7)** | | 37.8 | | (-8.0, 83.6) | |
|  | Routine and manual | **4.8** | | **(3.3, 6.2)** | | **97.2** | | **(57.2, 137.2)** | |
| Survey year ^IV^  (survey wave for data collection) | (1) 2008-2009 | Reference | |  | | Reference | |  | |
|  | (2) 2009-2010 | 0.8 | | (-2.0, 3.6) | | 23.3 | | (-56.4, 103.0) | |
|  | (3) 2010-2011 | 0.9 | | (-1.7, 3.5) | | 3.2 | | (-85.8, 92.1) | |
|  | (4) 2011-2012 | 0.2 | | (-2.8, 3.1) | | -54.9 | | (-139.55, 29.6) | |
|  | (5) 2012-2013 | -0.3 | | (-3.1, 2.6) | | -42.4 | | (-122.1, 37.2) | |
|  | (6) 2013-2014 | -0.4 | | (-3.5, 2.5) | | -60.2 | | (-137.7, 17.4) | |
|  | (7) 2014-2015 | 0.0 | | (-2.8, 2.8) | | -28.5 | | (-119.1, 62.0) | |
|  | (8) 2015-2016 | **-5.1** | | **(-8.2, -2.1)** | | **-134.3** | | **(-218.1, -50.6)** | |
|  | (9) 2016-2017 | **-4.1** | | **(-7.4, -0.8)** | | **-175.0** | | **(-259.0, -91.0)** | |
|  | (10) 2017-2018 | **-4.3** | | **(-7.6, -1.0)** | | -91.1 | | (-183.5, 1.2) | |
|  | (11) 2018-2019 | -2.7 | | (-6.0, 0.6) | | **-126.5** | | **(-219.2, -33.9)** | |
| Weight category^c^ | Normal weight | Reference | |  | | Reference | |  | |
|  | Overweight | 0.4 | | (-1.3, 2.2) | | 43.0 | | (-7.4, 93.3) | |
|  | Obese | 0.5 | | (-1.1, 2.1) | | **124.0** | | **(78.6, 169.4)** | |
| Ethnic group | White | Reference | |  | | Reference | |  | |
|  | Non-white | **-7.7** | | **(-9.6, -5.9)** | | **-199.4** | | **(-238.2, -160.6)** | |
| Region | England North | Reference | |  | | Reference | |  | |
|  | England Central/Midlands | -0.8 | | (-2.7, 1.1) | | 5.9 | | (-52.9, 64.7 | |
|  | England South (including London) | **-3.3** | | **(-5.0, -1.5)** | | **-105.2** | | **(-152.5, -58.0)** | |
|  | Scotland | 0.3 | | (-2.1, 2.7) | | -15.8 | | (-84.6, 52.9) | |
|  | Wales | -0.1 | | (-2.1, 2.0) | | -22.8 | | (-83.4, 37.8) | |
|  | Northern Ireland | 0.5 | | (-1.3, 2.3) | | **-73.5** | | **(-125.7, -21.3)** | |
| MVPA^d^ | <21 min/day | Reference | |  | | Reference | |  | |
|  | 21-52 min/day | 0.6 | | (-4.1, 5.2) | | 55.1 | | (-87.4, 180.4) | |
|  | 52 -124 min/day | 0.3 | | (-4.4, 5.0) | | -71.0 | | (-175.8, 103.6) | |
|  | >124 min/day | 0.4 | | (-4.7, 5.6) | | 22.5 | | (-96.5, 139.5) | |
| *Adjusted for Age, Sex, Survey Year and total energy intake (kcal/day) // **Only adjusted for Age and Survey Year when Sex was the exposure // ***Only adjusted for Sex and Survey Year when Age was the exposure  ^IV^ Only adjusted for Sex and Age when Survey year was the exposure.  ^a^ Results are weighed based on non-selection and non-response survey weights provided by NDNS year 2008-2019.  ^b^ Parents occupation is based on the three-class NS-SEC. A small number of households were excluded from this classification where the household has never worked or was classified under other.  ^c^ BMI z-score was created by standardising BMI for sex and age based on the 1990 British Growth Reference (UK90).  ^d^ MVPA: There was data available only for 16–18-year-olds, n=534.  Abbreviations: BMI z-score: standardised body mass index; CI: Confidence Interval; MVPA: Moderate-to-vigorous physical activity; NDNS: National Diet and Nutrition Survey; NS-SEC: National Statistics Socioeconomic Class; UPF: ultra-processed food, NOVA4 classification group.  Abbreviations: BMI z-score: standardised body mass index; MVPA: Moderate-to-vigorous physical activity; NDNS: National Diet and Nutrition Survey; NS-SEC: National Statistics Socio-economic Class; UPF: ultra-processed food, NOVA4 classification group. | | | | | | | | | |
